# Suicide among older nursing home residents in Austria: A nationwide register-based cohort study

**DOI:** 10.64898/2026.07.11.26357816

**Authors:** Erwin Stolz, Anna Schultz, Emilise Pötz, Carlos Watzka, Christian Jagsch, Annette Erlangsen

## Abstract

Relatively little is known regarding suicide among older adults in nursing homes. The aim of this study was to compare the incidence of suicide among older nursing home residents (NHR) with community-dwelling older people (CDP) using newly available, national, individual-level register data, and to assess differences with regard to socio-demographic characteristics. We obtained data on all older adults aged 65+ who were living in Austria at the end of October 2018 (n=1,665,450), including 155,020 NHR. Death by suicide was followed until the end of 2023. A total of 114 and 2,136 suicides were observed among NHR and CDP; corresponding to cumulative incidences of 14 and 27 per 100,000, respectively. Among NHR, suicide incidence was higher among males (28.0, 95% CI=22.1, 35.5), those aged 65-74 years (20.2, 95% CI=13.3, 30.6), with tertiary education (23.3, 95% CI=10.6, 50.6), divorced (25.0, 95% CI=16.2, 38.5), and residing in urban nursing homes (22.0, 95% CI=17.0, 28.4). Compared to CDP, more suicides in NHR occurred by poisoning and but few by firearms. In conclusion, we found that suicide incidence was lower among older NHR compared to CDP. More research on and preventive efforts against suicide among older NHR are needed.

## INTRODUCTION

Suicide rates are highest among the oldest old. Older adults aged 80 years and over have a two-fold higher suicide rate when compared to the total population^1^, and the suicide rates among older adults aged 80+ is particularly elevated in Austria (40/100,000)^a^.

Relatively little is known about suicide rates in different segments of older adults, such as nursing home residents (NHR).^2^ Findings from systematic reviews^3,4^ reveal that suicide ideation is common and suicide deaths are rare among NHR, and that the characteristics of NHR who die by suicide resemble those of older community-dwelling people (CDP).

However, these reviews are based on data from a small number of countries and few suicide deaths. The generalisability is therefore limited and more evidence is needed. Further, evidence regarding suicide incidences between NHR and CDP, and socio-demographic sub-groups thereof, may serve to inform preventive efforts^5^. The aim of this brief report was to assess suicide risk among older NHR in Austria.

## METHODS

A cohort design was applied to national data on all older adults living in Austria aged 65 years and over on 31^st^ of October 2018 (n=1,665,450). This group was followed until 31^st^ of December 2023. Individual-level data from the Central Residence Register and Central Civil Status Register were linked via a pseudonymised identifier.

Coroners, pathologists, and forensic pathologists determine causes of death in Austria. Suicide deaths were identified in the Central Civil Status Register using following codes from the 10^th^ revision of the International Classification of Diseases (ICD-10): X60-X84, Y87.0.

To avoid potential under-reporting, we included deaths with unclear intent (ICD-10: Y10-Y34) in a sensitivity analysis.

Socio-demographic markers included age (65-74, 75-84, ≥85 years), sex (males, females), level of education based on the International Classification of Education (primary, secondary, tertiary), and level of urbanisation (urban, town, rural). Information on mental disorders (ICD-10: F00-F99) was derived from the death certificates in Central Civil Status Register.

We calculated the annual cumulative incidence by dividing the number of suicides by the population at risk at the start of the observation period for NHR and CDP in total, and by age, sex, education and area of residence. Based on these, we subsequently calculated the cumulative incidence ratio (CIR) between NHR and CDP.

## RESULTS

Among the older adults aged 65 years, 63,223 lived in nursing home already at the beginning of the study. Another 91,797 individuals relocated to a nursing home during follow-up based, i.e. were recorded as living or having died in a nursing home. During the 5 years of follow-up, a total of 2,250 suicide were recorded. Of these, 114 (5.1%) occurred among NHR and 2,136 (9.3%) suicides among CDP, corresponding to annual cumulative incidences of 14 and 27 suicides per 100,000, respectively. The cumulative incidence ratio for NHR was 0.50 (95%-CI: 0.43, 0.62), suggesting a 50% lower suicide incidence among NHR. Suicide deaths comprised to 0.1% of all deaths among NHR. Among NHR, the most frequent methods of suicide were jumping (36.0%), hanging (28.1%), poisoning (18.4%), whereas hanging (38.6%), use of firearm (30.0%), and jumping (8.7%) were most frequent among CDP. When including deaths with unclear intent (n=560) in the sensitivity analysis, this increased incidences to 19 and 34 suicides per 100,000 for NHR and CDP, respectively, but did not alter0020the ratio substantively (CIR=0.57, 95%-CI=0.49, 0.57). The ratio of self-harm deaths with unclear intent compared to those with clear intent (i.e., suicides) was slightly higher in NHR (0.36) than in CDP (0.24).

With regard to socio-demographic characteristics, males had 3- and 5-fold higher cumulative incidences versus females among NHR and CDP, respectively (Table 1). The highest suicide incidences among NHR were found among those with higher levels of education and those divorced. The median age at death among NHR dying by suicide was 83 (IQR=79-88) years versus 79 (IQR=73-84) among CDP. Female NHR did seemingly not have lower cumulative incidences than female CDP. The difference between NHR and CDP was smallest for those aged 65-74 years (CIR=0.97; 95% CI=0.64, 1.47), suggesting equally high suicide incidences among the two groups. The difference was most pronounced among those aged 85 years and over (CIR=0.21, 95% CI=0.15, 0.29). NHR in urban areas had comparable suicide incidences as CDP (CIR=1.02, 95% CI=0.79, 1.32), whereas significantly lower suicide incidences were found among NHR in towns (CIR=0.31, 95% CI=0.20, 0.46) and rural areas (CIR=0.39, 95% CI=0.28, 0.55) versus CDP. Among those who died by suicide, 24.6% of NHR had been diagnosed mental disorder, while it was 22.9% of CDP. Most often, these were affective disorders (17.5% in NHR and 19.9% in CDP).

**Table 1:** Suicide deaths, cumulative incidence and incidence ratio among older nursing home residents and community-dwelling persons by socio-demographic characteristics ^a^ masked due prevent identifiability because of small numbers. CDR=community-dwelling resident, NHR=nursing-home resident, CI=confidence interval

|  | NHR | CDP | NHR | CDP |  |
| --- | --- | --- | --- | --- | --- |
|  | N (%) | N (%) | Incidence (95% CI) | Incidence (95% CI) | Incidence rate ratio (95% CI) |
| Total | 114 (5.1) | 2136 (94.9) | 14.2 (11.9, 17.1) | 27.4 (26.3, 28.6) | 0.50 (0.43, 0.62) |
| Sex |  |  |  |  |  |
| - Men | 68 (3.8) | 1706 (96.2) | 28.0 (22.1, 35.5) | 49.1 (46.8, 51.5) | 0.57 (0.45, 0.72) |
| - Women | 46 (9.7) | 430 (90.3) | 8.3 (6.18, 11.0) | 10.0 (9.06, 10.94) | 0.83 (0.62, 1.11) |
| Age |  |  |  |  |  |
| - 65-74 | 22 (2.5) | 860 (97.5) | 20.2 (13.3, 30.6) | 20.8 (19.5, 22.3) | 0.97 (0.64, 1.47) |
| - 75-84 | 58 (5.9) | 933 (94.2) | 18.9 (14.6, 24.4) | 32.7 (30.7, 34.9) | 0.58 (0.45, 0.75) |
| - 85+ | 34 (9.0) | 343 (91.0) | 8.8 (6.33, 12.35) | 42.3 (38.0, 47.0) | 0.21 (0.15, 0.29) |
| Education |  |  |  |  |  |
| - primary | 45 (6.7) | 628 (93.3) | 10.7 (8.01, 14.33) | 23.9 (22.1, 25.8) | 0.45 (0.34, 0.60) |
| - secondary | <65 (<4.5)<sup>a</sup> | 1360 (95.6) | 17.8 (13.9, 22.7) | 30.2 (28.7, 31.9) | 0.59 (0.46, 0.75) |
| - tertiary | <10 (<4.0)<sup>a</sup> | 148 (96.1) | 23.2 (10.6, 50.6) | 22.2 (18.9, 26.1) | 1.05 (0.47, 2.33) |
| Area of living |  |  |  |  |  |
| - Urban | 57 (9.9) | 519 (90.1) | 22.0 (17.0, 28.4) | 21.5 (19.8, 23.5) | 1.02 (0.79, 1.32) |
| - Town | 23 (3.3) | 675 (96.7) | 8.5 (5.69, 12.82) | 27.8 (25.8, 30.0) | 0.31 (0.20, 0.46) |
| - Rural | 34 (3.5) | 942 (96.5) | 12.5 (8.96, 17.49) | 31.9 (29.9, 34.0) | 0.39 (0.28, 0.55) |
| Marital status |  |  |  |  |  |
| - Married | 34 (2.7) | 1212 (97.3) | 16.1 (11.5, 22.5) | 26.0 (24.6, 27.5) | 0.62 (0.44, 0.87) |
| - Single | 14 (8.1) | 160 (92.0) | 16.6 (9.9, 27.9) | 36.5 (31.3, 42.6) | 0.46 (0.27, 0.77) |
| - Widowed | 46 (8.5) | 498 (91.5) | 10.8 (8.1, 14.4) | 25.9 (23.8, 28.3) | 0.42 (0.31, 0.56) |
| - Divorced | 20 (7.0) | 266 (93.0) | 25.0 (16.2, 38.5) | 34.2 (30.3, 38.6) | 0.73 (0.47, 1.13) |

## DISCUSSION

Using nationwide register data, the incidence of suicide among NHR was estimated for the first time in Austria. We found overall lower incidences for NHR than for CDP.

Given that the vast majority of older adults in Austria are CDP, this group drives the overall suicide rate.^1^ The national incidence of suicide among NHR in Austria was higher than in Australia^6^ but comparable to previous estimates from the US^7^ and Italy^8^. The rate of death by suicide among NHR has previously been found to be either higher^8,9^ or comparable^7^ with those of CDP. The seemingly protective or selection effect of being NHR in our study, might be explained through (1) a high proportion of females who have a lower suicide rate;^1^ (2) a high prevalence of physical impairment and neurodegenerative diseases, such as dementia, which have been linked to lower long-term risks of suicide;^10^ (3) close monitoring and care; (4) less easy access to means of suicide; and (5) those perceiving it as distressing to be a NHR might carry out suicidal acts prior to being relocated to a nursing home, i.e. while still being CDP.^7,9^ Due to lack of data, we were unable to confirm whether there was a higher rate of suicidal acts, as measured by suicide attempts before and after nursing home admission^9^ in Austria.

The 65–74-year-old NHR might represent a select group of individuals admitted to nursing homes at an unexpected age, possibly due to a specific debilitating reason, for instance substance misuse or disorders related to accelerated ageing, such as dementia. Hence, younger NHR residents might constitute an important target for prevention. Why higher education is linked to a higher suicide incidence among NHR but not CDP might relate to a larger perceived loss of autonomy in this group. The finding that one in four of NHR who died by suicide had been diagnosed mental health conditions and specifically affective disorders corresponds with existing evidence.^2–4,6,8^ Similar to earlier findings^3,4,9^, suicide by firearm was less common among NHR versus CDP. Nevertheless, it is noteworthy – and an option for intervention - that 6% of suicides in Austrian nursing homes were still by firearms.

Strengths of this analysis are the use of newly available, nationwide register data with individual-linkage. Limitations include that data was available only annually, i.e. it was not possible to assess temporal effects of nursing home admission and, further, had to calculate cumulative incidences instead of suicide rates, and hence our estimates are likely to be conservative. A low autopsy rate in Austria (7%), implies that some suicides deaths, particularly among NHR might have gone undetected as the rate of deaths due to self-harm with unclear intent was somewhat higher, although our sensitivity analysis supported our main findings. Finally, information on diagnoses, quality and amount of care, facility size and other related metrics^7^ was not available, which limited comparison to findings from other countries.^9^

In conclusion, we found that the cumulative incidence of suicide was lower among older NHR compared to CDP. Our analyses revealed distinct differences between the two groups with regard to age – suicide rate was highest among NHR aged 65-74 years and among CDP aged 85+ years – and suicide methods: poisoning was more and use of firearms was less prevalent among NHR compared to CDP.

## Patient consent for publication

Not applicable.

## Ethics approval

The conductance of this study was approved by the Ethics Committee of the Medical University of Graz (1172/2024).

## Data

Access and linkage of register data Central Residence Register, Central Civil Status Register, and Register-based Labour Market Statistics) was approved of and provided by the Austrian Micro Data Center (AMDC) of Statistics Austria following national legislation (Bundesstatistikgesetz §31, Forschungsorganisationsgesetz §38b). The AMDC is a research data infrastructure facility of Statistics Austria that enables research on micro data processed in compliance with data protection regulations. The data used for this research can be accessed by researchers at scientific institutions accredited with the AMDC against a fee. For further information, see https://www.statistik.at/en/services/tools/services/center-for-science/austrian-micro-data-center-amdc

## Footnotes

a https://www.sozialministerium.gv.at/dam/jcr:266e2eef-0a0a-4e30-95eb-b68142b6a95a/Suizidbericht%202025.pdf

